# What does simple power law kinetics tell about our response to coronavirus pandemic?

**DOI:** 10.1101/2020.04.03.20051797

**Authors:** Prateek K. Jha

## Abstract

Coronavirus pandemic of 2019-2020 has already affected over a million people and caused over 50,000 deaths worldwide (as on April 3, 2020). Roughly half of the world population has been asked to work from home and practice social distancing as the search for a vaccine continues. Though government interventions such as lockdown and social distancing are theoretically useful, its debatable whether such interventions are effective in flattening the curve, which is ceasing or reducing the growth of infection in control populations. In this article, I present a simple power law model that enables a comparison of countries in time windows of 14 days since first coronavirus related death is reported in that country. It therefore provides means to access the efficacy of above interventions.

## Introduction

Several epidemiological and statistical models[1–5] have been employed recently for the description of the coronavirus pandemic. Since the pandemic control is an ongoing effort, many of these models provide either an optimistic or a pessimistic picture. Nonetheless, such models are useful for government bodies to plan interventions and other measures to curb the disease. For instance, air travel restrictions between most affected countries came into existence by the middle of March’2020. Inbounds travelers from high risk countries usually went through thermal screening at the airport, followed by strict isolation of symptomatic and quarantine for asymptomatic travelers. Further, most countries have adopted strategies to encourage social distancing in several phases, e.g., (1) educating citizens to avoid physical contacts, (2) limiting social gathering, (3) closing academic institutions, (4) strict lockdown with only essential services open. Finally, many countries have adopted random testing in populations to identify affected people.

To the best of my knowledge of the scientific understanding of coronavirus at the point of writing this article,

- Infection mainly occurs through physical contact and is not airborne.
- The effect of weather on the spread of pandemic has not been established.
- Although the older people have high death risk due to comorbities, the virus also affects younger populations. In fact, asymptomatic young people may be silent virus carriers.
- Travel restrictions between countries are very useful in the initial phases of the pandemic but have little effect in the later phases.

Epidemiological models can be developed with the objective of either understanding the effect of various interventions on the spread of the infection or to analyze and forecast the magnitude of infections or death in different geographical regions. One of the simplest and intuitive models can be developed in the following way. Lets consider *i =* 1, *2*,…. *N* regions of a system containing 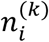 active infections on day *k*. The system can be world and the regions can be countries, or the system can be a country and the regions can be states, or the system can be a state and the regions can be cities, etc.

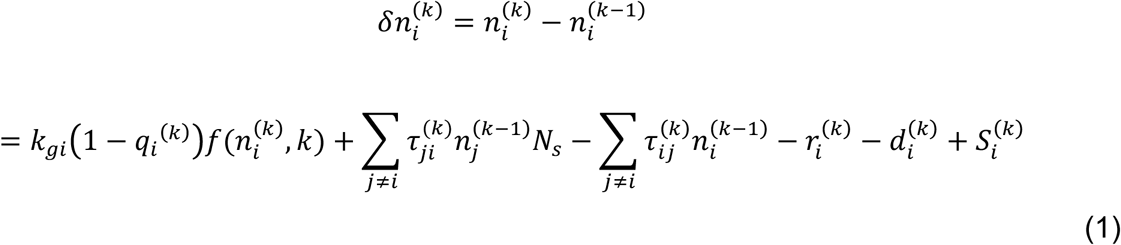

Here, *k*_*gi*_ is the rate constant for spread of infection from existing cases in region *i*. 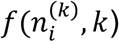 is an arbitrary function that captures the nature of spread. For example, 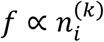 for a first-order process. *q*_*i*_^(*k*)^ is the fraction of new cases that were quarantined in region *i* on day *k*, where we assume that the quarantined cases do not infect further. 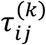 is the fraction of infected people of region *i* going to region *j*, who have not been tested. *N*_*s*_ is the number of secondary infections caused by an infected person during the journey, e.g. at the airport. 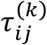 can be approximated as 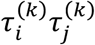, where 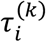 is the fraction of infected case traveling to other regions. 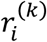 and 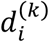 are the number of cases recovered and died on day 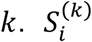 is the number of new cases coming from outside the system (if system is not world) or emerging within system. Unfortunately, eq. (1) contains many parameters that are difficult to estimate in the middle of a pandemic as the pandemic has not properly evolved in many regions. Nevertheless, the beneficial effects of quarantine and travel restrictions can be easily seen from eq. (1). In a hypothetical situation of full quarantine 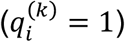 and no travel 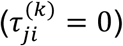, the growth is completely ceased unless new cases emerge within the region 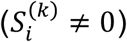. In realistic situations, none of the above condition can be fully met. However, if 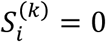 and *f*grows with *k* more rapidly than the first-order growth in second term of eq. (1), the functional form of *f* in eq. (1) dictates the growth behavior.

I have tried several functions to fit the growth behavior and observed that power law functions of the type 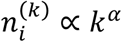 with different exponents at different time windows approximately captures the growth behavior of almost all countries. Similar fractal scaling has been discussed by Ziff in a recent work[1]. Any serious intervention is expected to ‘flatten’ the curve by reducing *α*. The details of our methodology and obtained results are discussed in the following sections.

## Methodology

I have used the Covid data set of Johns Hopkins University (https://coronavirus.jhu.edu/), which contained data until April 1, 2020 at the time of download. Although the fitting was performed for confirmed cases, recovered cases, and deaths per day, I only report the results for deaths per day. This is because the number of reported confirmed cases may significantly depart from the actual number of confirmed cases due to a systematic lack of population-wide testing and the infection may reappear in asymptomatic and recovered cases. Countries with atleast 10 reported deaths until April 1, 2020, are only considered henceforth. For each country, Day 0 is determined as the day when the first death case is reported. Following power-law relation is fitted,

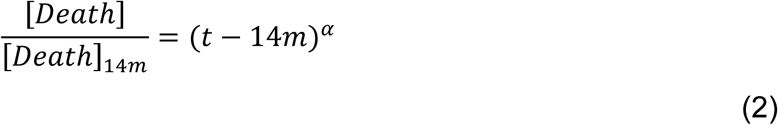

where separate exponent *α* is computed for 0-14 days (*m*=0), 14-28 days (*m*= 1), 28-42 days (*m*= *2*), etc. Mean *α*_*m*_ and standard deviation *σ*_*m*_ of *α* values are computed. Here, *m* represents the number of countries for which the time window is applicable. For instance, if Day 0 of a country is 20 days prior to April 1, 2020, it will be considered in the time windows of 0-14 days and 14-28 days but not considered in 28-42 days, 42-56 days, etc. Following risk-levels are assigned for countries in each time window:

- SAFE if *α*<*α*_*m*_−*σ*_*m*_
- MODERATE if *α*_*m*_−*σ*_*m*_≤*α*<*α*_*m*_
- HIGH if *α*_*m*_≤*α*<*α*_*m*_+*σ*_*m*_
- DANGEROUS if *α*≥*α*_*m*_+*σ*_*m*_

## Results and Discussion

Table 1 shows the mean and standard deviation of *α* in different time windows averaged over the countries for which the first death has occurred before the beginning of the time window. As expected, *α*_*m*_ decreases with passage of time and the pandemic can be assumed to be controlled after 70 days. Note however that while the exponents decrease, the number of cases still remain high and the actual pandemic control would depend on the recovery rate of infected individuals. The standard deviation *σ*_*m*_ is almost half of *α*_*m*_ until 28 days but become comparable to *α*_*m*_ beyond this period, which may be attributed to the smaller number of countries over which averaging is performed.

**Table 1:** Mean and standard deviation in time windows.

| Time Window | Number of countries | $\alpha_m$ | $\sigma_m$ |
| --- | --- | --- | --- |
| **0-14** | 65 | 1.0 | 0.5 |
| **14-28** | 42 | 0.5 | 0.3 |
| **28-42** | 11 | 0.4 | 0.3 |
| **42-56** | 4 | 0.2 | 0.2 |
| **56-70** | 2 | 0.1 | 0.1 |

Figure 1 shows the predicted number of deaths using *α*_*m*_ from Table 1 (bold line) and the optimistic and pessimistic scenarios corresponding to *α*_*m*_±*σ*_*m*_ (dashed lines). The actual death data of some representative countries are shown in the figure. The efficiency of a country in controlling the pandemic can be accessed by determining risk levels. Note that if a country is in the DANGEROUS level in a time window, it will not immediately go to regions of lower risk in Figure 1 even when *α* drops. Therefore, in order to contain the pandemic, a country should begin interventions in the first time window. Interventions occurring in later time windows have lesser effect also because they do not affect the already infected cases that have been not detected or were asymptomatic. Table 2 summarizes the country-wise statistics where the DANGEROUS time windows of countries are colored.

**Table 2:**
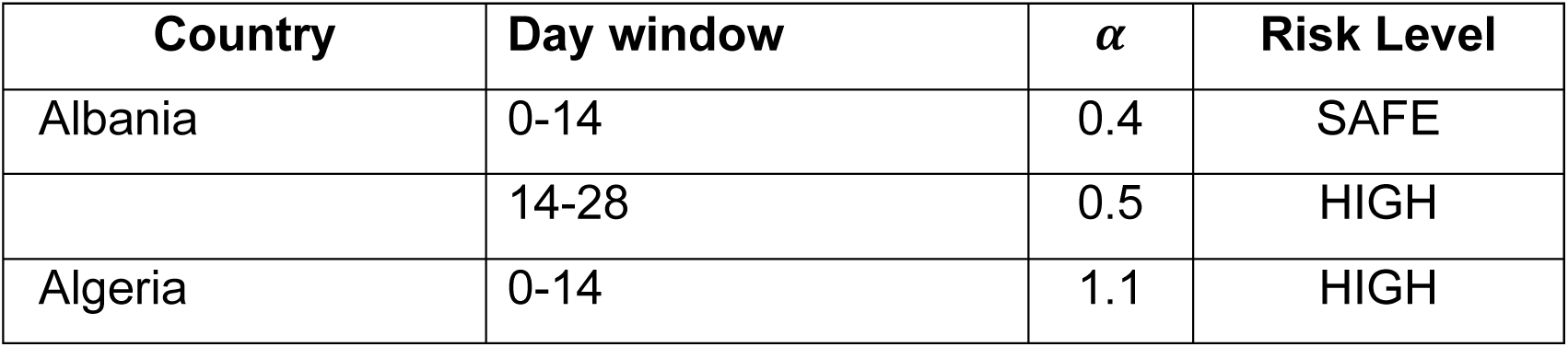
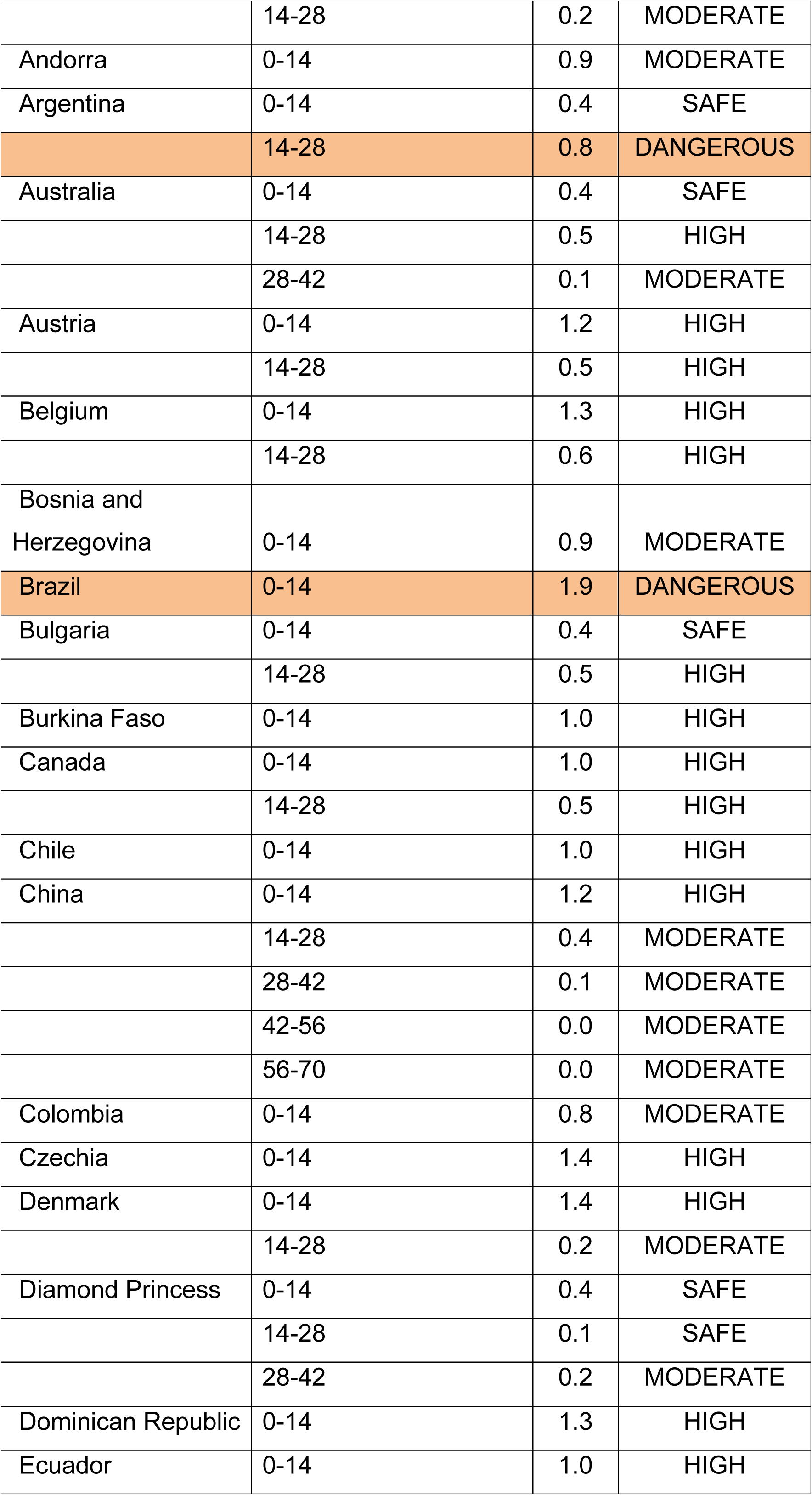
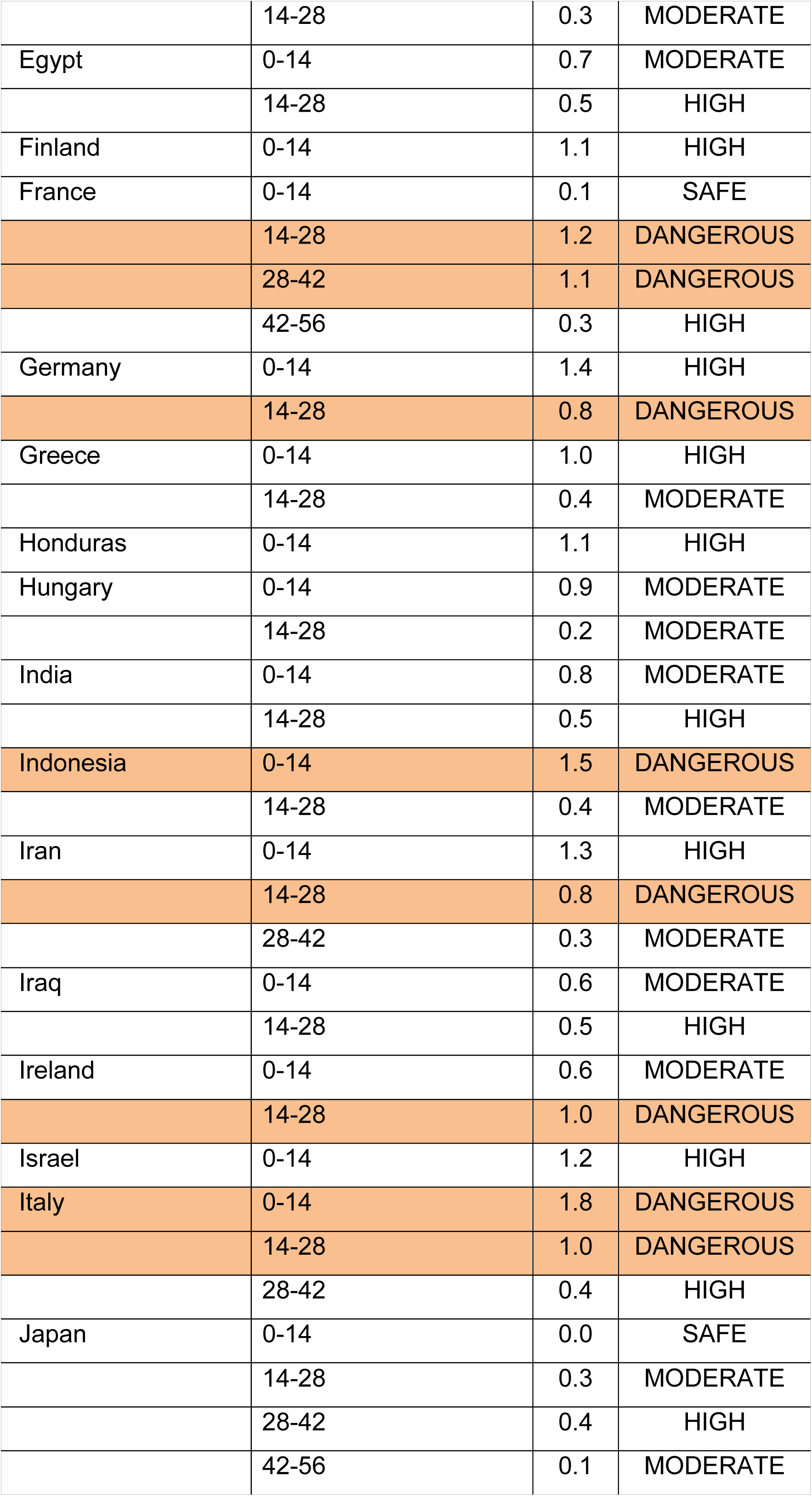
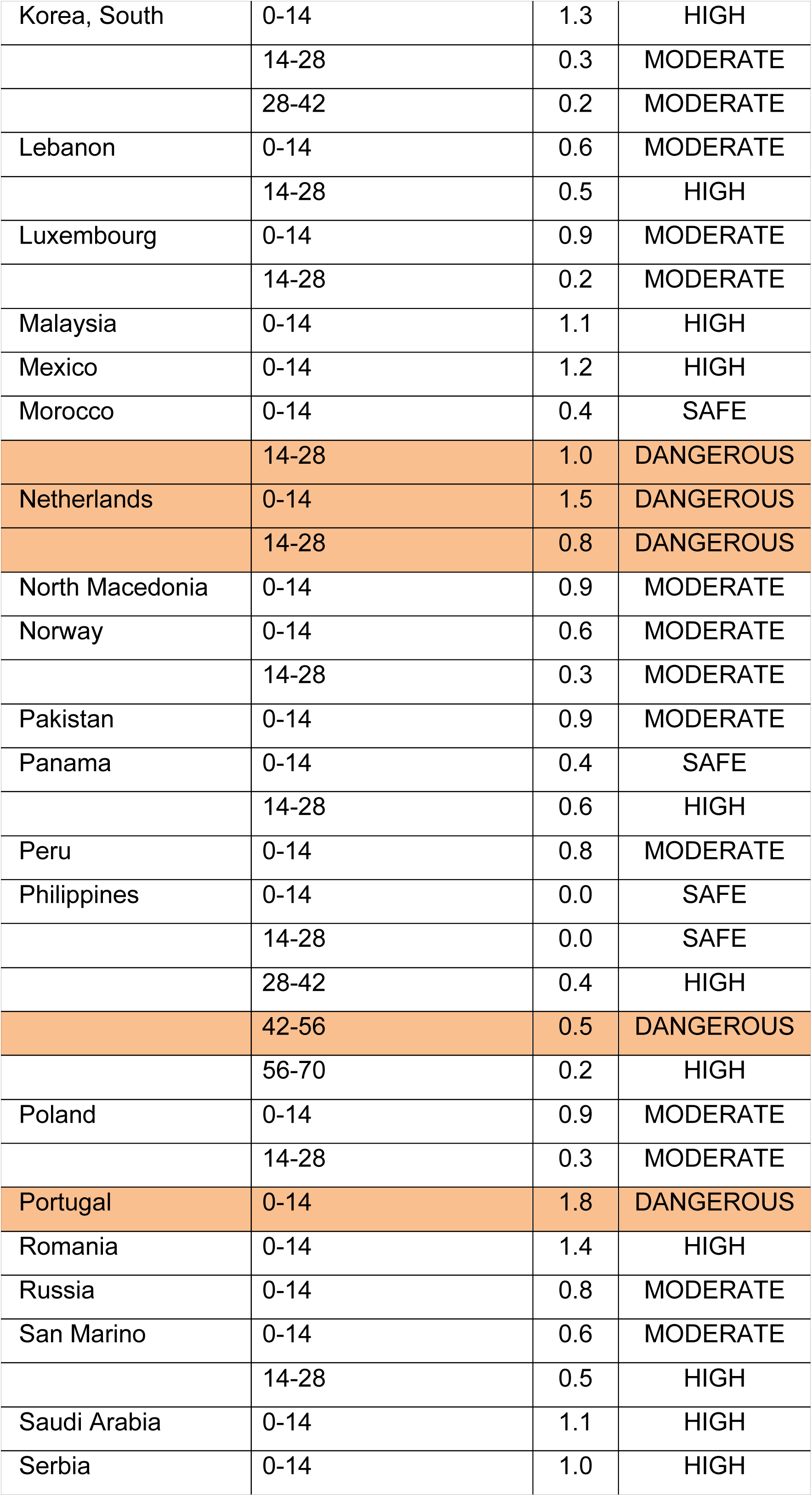
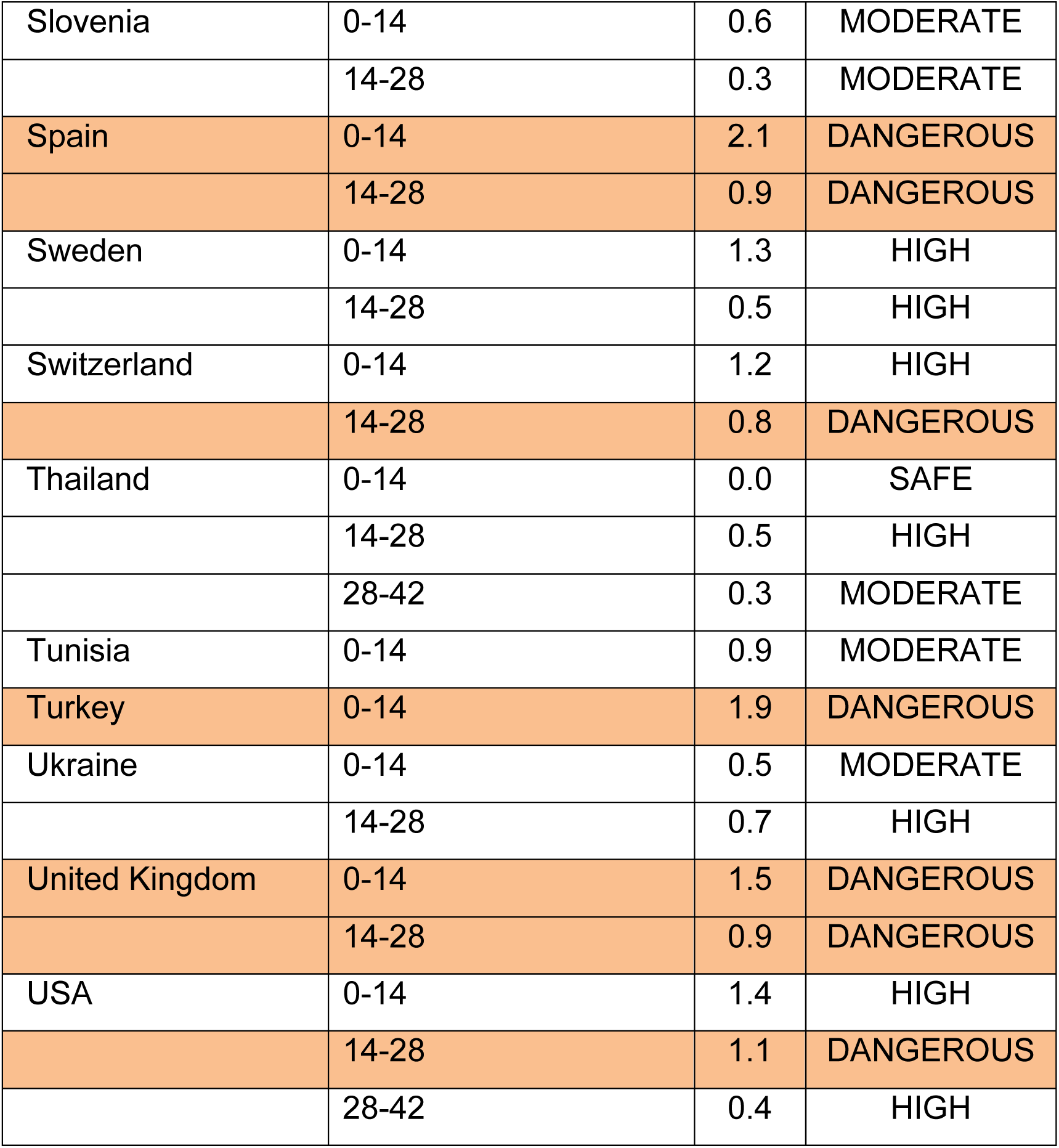
Country-wise values of *α* in eq. (2) that fit the death data. “DANGEROUS” risk levels are colored in the table. See text for description of risk levels.

| Country | Day window | $\alpha$ | Risk Level |
| --- | --- | --- | --- |
| Albania | 0-14 | 0.4 | SAFE |
|  | 14-28 | 0.5 | HIGH |
| Algeria | 0-14 | 1.1 | HIGH |
|  | 14-28 | 0.2 | MODERATE |
| Andorra | 0-14 | 0.9 | MODERATE |
| Argentina | 0-14 | 0.4 | SAFE |
|  | 14-28 | 0.8 | DANGEROUS |
| Australia | 0-14 | 0.4 | SAFE |
|  | 14-28 | 0.5 | HIGH |
|  | 28-42 | 0.1 | MODERATE |
| Austria | 0-14 | 1.2 | HIGH |
|  | 14-28 | 0.5 | HIGH |
| Belgium | 0-14 | 1.3 | HIGH |
|  | 14-28 | 0.6 | HIGH |
| Bosnia and Herzegovina | 0-14 | 0.9 | MODERATE |
| Brazil | 0-14 | 1.9 | DANGEROUS |
| Bulgaria | 0-14 | 0.4 | SAFE |
|  | 14-28 | 0.5 | HIGH |
| Burkina Faso | 0-14 | 1.0 | HIGH |
| Canada | 0-14 | 1.0 | HIGH |
|  | 14-28 | 0.5 | HIGH |
| Chile | 0-14 | 1.0 | HIGH |
| China | 0-14 | 1.2 | HIGH |
|  | 14-28 | 0.4 | MODERATE |
|  | 28-42 | 0.1 | MODERATE |
|  | 42-56 | 0.0 | MODERATE |
|  | 56-70 | 0.0 | MODERATE |
| Colombia | 0-14 | 0.8 | MODERATE |
| Czechia | 0-14 | 1.4 | HIGH |
| Denmark | 0-14 | 1.4 | HIGH |
|  | 14-28 | 0.2 | MODERATE |
| Diamond Princess | 0-14 | 0.4 | SAFE |
|  | 14-28 | 0.1 | SAFE |
|  | 28-42 | 0.2 | MODERATE |
| Dominican Republic | 0-14 | 1.3 | HIGH |
| Ecuador | 0-14 | 1.0 | HIGH |
|  | 14-28 | 0.3 | MODERATE |
| Egypt | 0-14 | 0.7 | MODERATE |
|  | 14-28 | 0.5 | HIGH |
| Finland | 0-14 | 1.1 | HIGH |
| France | 0-14 | 0.1 | SAFE |
|  | 14-28 | 1.2 | DANGEROUS |
|  | 28-42 | 1.1 | DANGEROUS |
|  | 42-56 | 0.3 | HIGH |
| Germany | 0-14 | 1.4 | HIGH |
|  | 14-28 | 0.8 | DANGEROUS |
| Greece | 0-14 | 1.0 | HIGH |
|  | 14-28 | 0.4 | MODERATE |
| Honduras | 0-14 | 1.1 | HIGH |
| Hungary | 0-14 | 0.9 | MODERATE |
|  | 14-28 | 0.2 | MODERATE |
| India | 0-14 | 0.8 | MODERATE |
|  | 14-28 | 0.5 | HIGH |
| Indonesia | 0-14 | 1.5 | DANGEROUS |
|  | 14-28 | 0.4 | MODERATE |
| Iran | 0-14 | 1.3 | HIGH |
|  | 14-28 | 0.8 | DANGEROUS |
|  | 28-42 | 0.3 | MODERATE |
| Iraq | 0-14 | 0.6 | MODERATE |
|  | 14-28 | 0.5 | HIGH |
| Ireland | 0-14 | 0.6 | MODERATE |
|  | 14-28 | 1.0 | DANGEROUS |
| Israel | 0-14 | 1.2 | HIGH |
| Italy | 0-14 | 1.8 | DANGEROUS |
|  | 14-28 | 1.0 | DANGEROUS |
|  | 28-42 | 0.4 | HIGH |
| Japan | 0-14 | 0.0 | SAFE |
|  | 14-28 | 0.3 | MODERATE |
|  | 28-42 | 0.4 | HIGH |
|  | 42-56 | 0.1 | MODERATE |
| Korea, South | 0-14 | 1.3 | HIGH |
|  | 14-28 | 0.3 | MODERATE |
|  | 28-42 | 0.2 | MODERATE |
| Lebanon | 0-14 | 0.6 | MODERATE |
|  | 14-28 | 0.5 | HIGH |
| Luxembourg | 0-14 | 0.9 | MODERATE |
|  | 14-28 | 0.2 | MODERATE |
| Malaysia | 0-14 | 1.1 | HIGH |
| Mexico | 0-14 | 1.2 | HIGH |
| Morocco | 0-14 | 0.4 | SAFE |
|  | 14-28 | 1.0 | DANGEROUS |
| Netherlands | 0-14 | 1.5 | DANGEROUS |
|  | 14-28 | 0.8 | DANGEROUS |
| North Macedonia | 0-14 | 0.9 | MODERATE |
| Norway | 0-14 | 0.6 | MODERATE |
|  | 14-28 | 0.3 | MODERATE |
| Pakistan | 0-14 | 0.9 | MODERATE |
| Panama | 0-14 | 0.4 | SAFE |
|  | 14-28 | 0.6 | HIGH |
| Peru | 0-14 | 0.8 | MODERATE |
| Philippines | 0-14 | 0.0 | SAFE |
|  | 14-28 | 0.0 | SAFE |
|  | 28-42 | 0.4 | HIGH |
|  | 42-56 | 0.5 | DANGEROUS |
|  | 56-70 | 0.2 | HIGH |
| Poland | 0-14 | 0.9 | MODERATE |
|  | 14-28 | 0.3 | MODERATE |
| Portugal | 0-14 | 1.8 | DANGEROUS |
| Romania | 0-14 | 1.4 | HIGH |
| Russia | 0-14 | 0.8 | MODERATE |
| San Marino | 0-14 | 0.6 | MODERATE |
|  | 14-28 | 0.5 | HIGH |
| Saudi Arabia | 0-14 | 1.1 | HIGH |
| Serbia | 0-14 | 1.0 | HIGH |
| Slovenia | 0-14 | 0.6 | MODERATE |
|  | 14-28 | 0.3 | MODERATE |
| Spain | 0-14 | 2.1 | DANGEROUS |
|  | 14-28 | 0.9 | DANGEROUS |
| Sweden | 0-14 | 1.3 | HIGH |
|  | 14-28 | 0.5 | HIGH |
| Switzerland | 0-14 | 1.2 | HIGH |
|  | 14-28 | 0.8 | DANGEROUS |
| Thailand | 0-14 | 0.0 | SAFE |
|  | 14-28 | 0.5 | HIGH |
|  | 28-42 | 0.3 | MODERATE |
| Tunisia | 0-14 | 0.9 | MODERATE |
| Turkey | 0-14 | 1.9 | DANGEROUS |
| Ukraine | 0-14 | 0.5 | MODERATE |
|  | 14-28 | 0.7 | HIGH |
| United Kingdom | 0-14 | 1.5 | DANGEROUS |
|  | 14-28 | 0.9 | DANGEROUS |
| USA | 0-14 | 1.4 | HIGH |
|  | 14-28 | 1.1 | DANGEROUS |
|  | 28-42 | 0.4 | HIGH |

**Figure 1:**
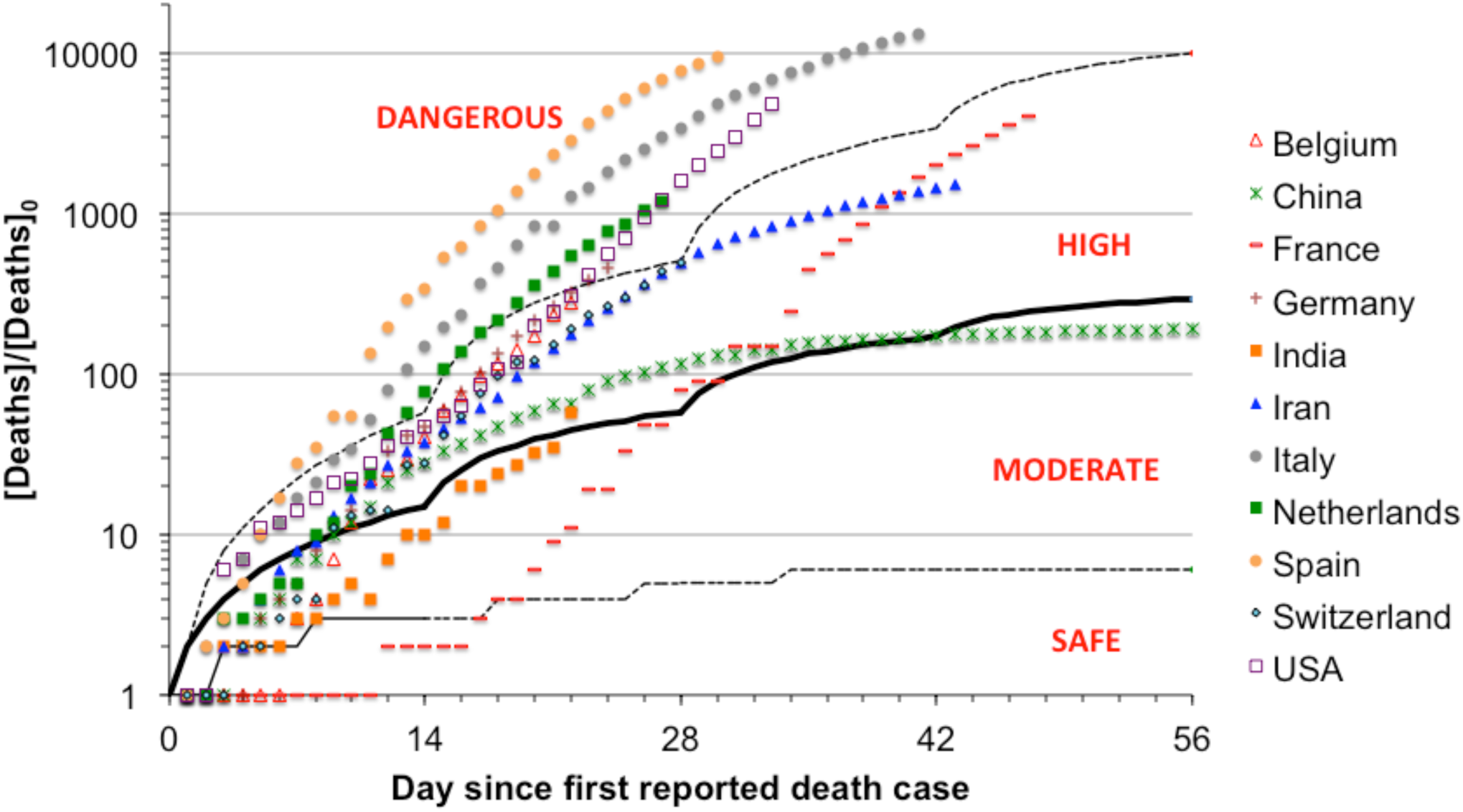
Total coronavirus 2019-2020 deaths for some of the countries, normalized by deaths on Day 0. Bold line indicate the one obtained by using *α*_*m*_ values in Table 1 in eq. (2). Dashed lines indicate the lines obtained by using *α*_*m*_±*σ*_*m*_ values in Table 1 in eq. (2). Different risk levels are indicated in figure.

## Conclusion

A simple power law model has been developed that analyzed the responses of countries to the Coronavirus 2019-2020 pandemic. The results of this study, although preliminary, is expected to provide useful insights on the ongoing efforts to contain the pandemic.

## Data Availability

Data can be obtained from the author on request.

## Acknowledgments

Author thanks Pankaj Doshi and Pulkit Sharma for discussions. The author himself was in quarantine and working from home, while writing this article.

